# Identification of Novel Gene variants for Autism Spectrum Disorder in an Indian Patient using Whole Exome Sequencing

**DOI:** 10.1101/2024.02.28.24303417

**Authors:** Prashasti Yadav, Saileyee Roychowdhury, Nilanjan Mukherjee, Reema Mukherjee, Sudipta Kumar Roy, Soumen Bhattacharjee, Parimal Das

## Abstract

**Background:** Autism Spectrum Disorder (ASD) is a neurodevelopmental disorder characterized by persistent deficits in social communication and interaction, along with restricted and repetitive behaviour patterns, interests or activities. Its prevalence has risen over the past few years, being four times more common in boys than girls. The cause of ASD is unclear, its etiology involves genetic, environmental, and gene-environment interactions. While past studies highlighted clinical genetic risks, genetic complexity of ASD, with variants of diverse frequencies, type, and inheritance patterns, requires further exploration for better management of disease. Researches have shown that the whole exome sequencing can be used to identify genetic variants associated with genetically heterogeneous conditions. The purpose of this study is to identify genetic variants by employing whole exome sequencing in an Indian ASD patient.

**Methods:** A female patient of age within 0-5 years, having characteristic features like hyperactivity and language impairment, was investigated and diagnosed using DSM-5 criteria. Peripheral blood sample collection was done followed by DNA extraction and whole exome sequencing. Variants analysis, identification and annotation were done using bioinformatics tools and databases. Identified pathogenic variants were reconfirmed by Sanger sequencing.

**Results and conclusion:** Our study uncover four genetic variations, comprising three missense variations in *KIF1A* (c.3839C>T), *SETD5* (c.314A>C), *MAPK81P3* (c.2849C>T), and one-stop gain variation in *ERMARD* (c.1523G>A). The *ERMARD* stop gain variation, predicted to induce nonsense-mediated decay, alter normal protein function through truncation and classified as likely pathogenic based on the ACMG guidelines and current available scientific evidence. In conclusion, we identified a likely pathogenic variant in *ERMARD* along with three missense variants in *KIF1A, SETD5* and *MAPK81P3* respectively. These findings suggest the potential contribution of *ERMARD* mutations to ASD susceptibility, emphasizing the need for further validation through functional studies.

## Introduction

Autism Spectrum Disorder (ASD) is a common heterogenous lifelong neurodevelopmental condition ranging from mild to severe and characterized by persistent deficits in social communication and social interaction, restricted, repetitive patterns of behaviour, interests, or activities with unusual sensory-motor functions [1, 2]. Due to the absence of reliable biomarkers, the diagnosis most often is based upon the behaviour of the child. In recent years, its prevalence has gradually increased and become four times more common among boys than girls [3]. The estimated global prevalence of ASD is one in 100 [4]. The reported prevalence of ASD in South Asia is estimated to be one in 93 [5]. In India, the estimated prevalence of ASD in rural areas is 0.11% while in urban areas it is 0.09% (ages 1-18 years) [6]. ASD is clinically heterogeneous, some individuals presenting mild symptoms and others experiencing severe symptoms with a range of co-occurring physical and mental health conditions [7]. The etiology of ASD has not been understood, studies have shown that it may be multifactorial; genes, environment and gene-environment interactions play important role in the pathogenesis [8]. Large number of genes reported to be involved in the pathogenesis ASD, majority of which expressed in neuronal cells and enriched in maturing neurons [9,10,11,12] and thought to converge on common pathways affecting neuronal and synaptic homeostasis [13]. Pathway network analyses of gene ontologies suggest that, genes contributing to the core features of ASD may also contribute to other vulnerabilities, that is important molecular mechanisms leading to multiple systemic comorbidities that also overlap with other conditions [14]. Variations in multiple genes show the strong evidence of involvement of genetic factor in the pathogenesis of ASD [15]. Twin studies suggest the heritability of ASD to be 64%-91% [16]. Previous studies show the chromosomal abnormalities, copy number variations (CNVs) and single nucleotide variations (SNV) have been associated with ASD [17]. Rare or *de novo* genetic variants are identified in 5%-20% of individuals with ASD, and more often associated with complex medical presentation [18]. Rare variants causing ASD risk collectively encompass hundreds of genes [19], while copy-number variant and *de novo* protein-altering mutations show extreme locus heterogeneity [20]. In recent years development in genomic sequencing have transformed variant discovery, different approaches have been used to discover the genetic variants associated with ASD. Whole exome sequencing has been used to identify rare and novel genetic variation related to neurodevelopmental disorders [21] and have greatly improved the chance of identifying known as well as novel responsible genes [22, 23]. Studies has reported the combining clinical and molecular diagnosis is fundamental to deepen the knowledge of the pathogenic mechanisms of neurodevelopmental disorders underlying medical conditions and to develop personalized treatments [24].

In the present study, WES was performed for a patient sample with a diagnosis of an ASD related phenotype. We identified four genetic variations, including three missense and one stop gain variation. We select the variant which are predicted to alter normal protein function through protein truncation and classified as likely pathogenic for the reported phenotype based on current available scientific evidence using ACMG guideline [25].

In conclusion findings of this study provide valuable insights for pathogenicity of genetic variations and shed light on the underlying molecular process involved in ASD and confirm the efficacy of WES in detecting pathogenic variants in ASD candidate genes.

## Material and Methods

### Recruitment of patient and sample collection

Patient was enrolled from West Bengal, India. The study protocol was approved by the Institutional Ethics Committee of Centre for Genetic Disorders, Institute of Science, Banaras Hindu University, Varanasi.

The diagnosis of ASD was done according to the American Psychiatric Association’s Diagnostic and Statistical Manual of Mental Disorders (DSM-5) criteria [1] and ICD-10 [International Classification of Diseases, Tenth Revision] [26], and also evaluation was done using standard scale, IASQ (The Indian Autism Screening Questionnaire) [27].

As the proband was minor, peripheral blood sample was collected after obtaining the written informed consent from parents.

### DNA Extraction and Whole exome sequencing

Genomic DNA was extracted using salting out method [28]. Whole exome sequencing was performed on genomic DNA sample of the proband. Sequencing of the protein coding regions of approximately 30 Mb of the human exome (targeting approximately 99% of the regions in CCDS and Refseq) was performed using Illumina next generation sequencing (NGS) systems at a mean depth of 50-60X and percentage of bases covered at 20X depth >90% in the target region.

### Variant filtration and identification

Alignment of obtained sequences to human reference genome (GRCh37/hg19) was done using BWA-mem aligner. Variant calling was obtained using Genome Analysis ToolKit (GATK). Duplicate reads identification and removal, Base quality recalibration and re-alignment of reads based on indels were done using inbuilt Sentieon modules [29]. Sention’s Haplotype caller module was used to identify the variants which were relevant to the clinical indications along with the Deep variant analysis pipeline on Google cloud platform which was used as a secondary pipeline to call genetic variants [30]. Quality checks (QC) were performed on all VCF files to exclude variants where sequencing was of poor quality. Additional QC metrics includes total homozygous and heterozygous calls (SNVs and indels), proportion of variant calls that were common, number of variants falling into different annotated consequence categories, number of extreme heterozygous (alternative allele proportion 0.8).

### Variant annotation and classification

The following public databases were used for annotation of identified variants: OMIM, GWAS, GNOMAD, 1000 Genomes database [31, 32, 33]. For the interpretation of variants, the American College of Medical Genetics and Genomics (ACMG) 2015 guidelines were used [25].

### Sanger sequencing

To confirm the *ERMARD* variant, we performed Sanger sequencing of exon 16, including the flanking intron sequences of the gene (NM_018341.3) in the proband. PCR was done with specific primer pairs to amplify DNA, followed by purification of the PCR product. Subsequently, the purified PCR product underwent Sanger sequencing using an ABI 3500 Genetic Analyzer (Applied Biosystems, USA) according to the manufacturer’s protocol. The Sanger sequencing results were analysed with sequence scanner software 2 v2.0. Visualization of DNA sequences were performed by Finch TV v1.4.0 (Geospiza, PerkinElmer, USA) software.

## Results

### Clinical Description

This study encompasses a single Indian family, with the proband being a female of age within 0-5 years. She exhibited severe hyperactivity, speech regression, and an inability to articulate meaningful words.

She has no family history of ASD or other neurodevelopmental disorders.

### WES Analysis

Whole exome sequencing data revealed a likely pathogenic stop-gain variant c.1523G>A in exon 16 of the *ERMARD* gene on chromosome 6 along with three missense variants; *KIF1A* (c.3839C>T), *SETD5* (c.314A>C), *MAPK81P3* (c.2849C>T), classified as variant of uncertain significance based on current available scientific evidence for the reported phenotype [Table 1].

**Table 1.** Findings related to phenotype.

| Gene & transcript | Variant | Location | Zygosity | Inheritance | Classification |
| --- | --- | --- | --- | --- | --- |
| *ERMARD*<br>NM_018341.3 | c.1523G>A<br>(p.Trp508Ter) | Exon 16 | Heterozygous | Autosomal<br>Dominant | Likely<br>Pathogenic |
| *KIF1A*<br>NM_004321.8 | c.3839C>T<br>(p.Pro1280Le) | Exon 38 | Heterozygous | Autosomal<br>Dominant | Uncertain<br>Significance |
| *SETD5*<br>NM_001080517.3 | c.314A>C<br>(p.Asn105Thr) | Exon 5 | Heterozygous | Autosomal<br>Dominant | Uncertain<br>Significance |
| *MAPK8IP3*<br>NM_015133.5 | c.2849C>T<br>(p.Thr950Ile) | Exon 23 | Heterozygous | Autosomal<br>Dominant | Uncertain<br>Significance |

Sanger sequencing confirmed the heterozygous *ERMARD* c.1523G>A variant in the proband [Figure 1]. This stop gain variant has not been previously reported and it is not present in gnomAD and 1000 genomes databases. This variation occurs upstream in exon 16 of *ERMARD* is predicted to be a nonsense mediated decay which alter normal protein function through protein truncation.

**Figure 1.**
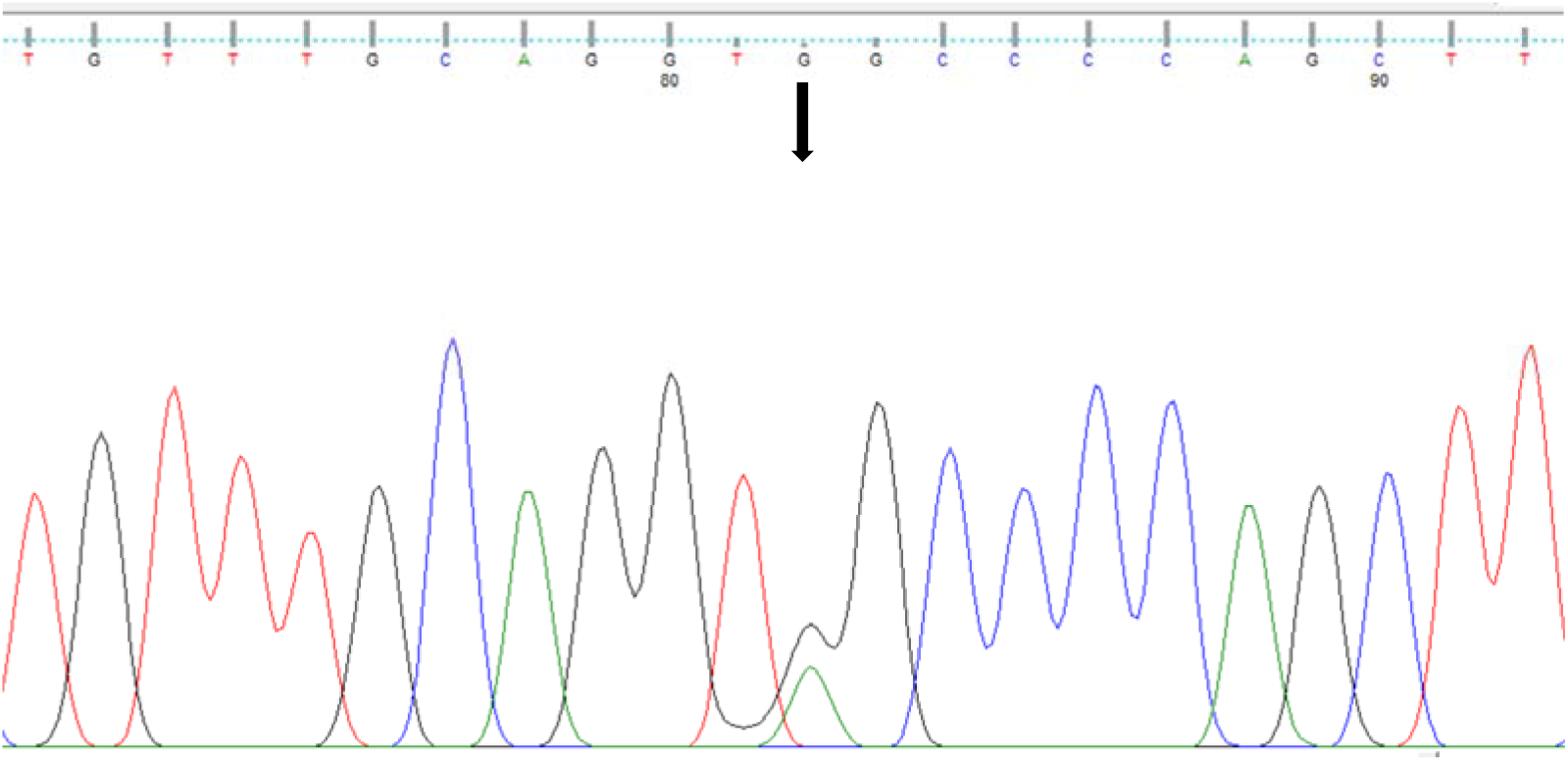
Confirmation of heterozygous variant c.1523G>A (denoted by arrow) by sanger sequencing.

## Discussion

In present study, we identified a likely pathogenic variant c.1523G>A of *ERMARD* gene along with three variants of uncertain significance in genes; *KIF1A* (c.3839C>T), *SETD5* (c.314A>C) and *MAPK8IP3* (c.2849C>T) for the reported phenotype of a patient having characteristic features of ASD. These variations have not been reported previously as a pathogenic or benign and also not present in gnomAD and 1000 genomes databases.

*ERMARD* (ER membrane associated RNA degradation) gene encodes a protein with two transmembrane domains near the C-terminus and localised in the endoplasmic reticulum. Also known as C6ORF70, it is present on chromosome 6q27 [34]. According to previous studies the heterozygous mutations in the *ERMARD* have been associated with Periventricular nodular heterotopia-6 (PVNH6), a disease characterized by delayed psychomotor development, delayed speech, strabismus, onset of seizures with hypsarrhythmia and brain MRI showing bilateral periventricular nodular heterotopia in the frontal horns [34].

*KIF1A* encodes a motor protein that is involved in the anterograde transport of synaptic vesicle precursors along axons [35]. Mutations in *KIF1A* have been associated with a wide range of conditions including recessive mutations causing hereditary sensory neuropathy and hereditary spastic paraplegia [35,36] and *de novo* dominant mutations causing intellectual disability, cerebellar atrophy, spastic paraparesis, optic nerve atrophy, peripheral neuropathy, and epilepsy [37]. A *de novo* dominant missense variant has been reported in a patient presenting with ASD, spastic paraplegia and axonal neuropathy [38].

*SETD5* is located on chromosome 3p25.3 and encodes the SETD5 protein composed of 1442 amino acids [39], and consists of 31 exons and is ubiquitously expressed in human tissues such as the brain, thyroid, skin, ovary, lung and endometrium [40, 41]. SETD5 contains a SET domain and is thus annotated as a candidate protein of lysine methyltransferase, which methylates H3K36 up to the tri-methyl form (H3K36me3) [40, 42, 43]. Autosomal dominant mental retardation-23 (MRD23) is caused by heterozygous mutation in the SETD5 gene, characterized by moderate to severe intellectual disability, delayed psychomotor development in infancy, poor speech development, obsessive-compulsive behaviour, hand-flapping and features of autism [44].

*MAPK8IP3* encodes a member of the kinesin superfamily of proteins and plays a role in axonal transport [45]. Heterozygous mutation in the *MAPK8IP3* caused neurodevelopmental disorder with or without variable brain abnormalities (NEDBA) [46].

In conclusion, findings of this study suggests that mutation in *ERMARD* with other reported variation in genes: *KIF1A, SETD5* and *MAPK8IP3* may cause the reported phenotype. Further functional studies in cell and animal models are needed to elucidate the role of variant in the pathogenesis of ASD.

## Data Availability

All data produced in the present work are contained in the manuscript

## Notes

### Competing Interest Statement

The authors have declared no competing interest.

### Funding Statement

The first author acknowledges Indian Council of Medical Research, India For providing Junior Research Fellowship (JRF) and Senior Research Fellowship (SRF)

### Author Declarations

Ethics Committee Institute of Science Banaras Hindu University Varanasi-221005, INDIA (Ref No: I.Sc./ECM-XVI/2023-2024/)

## References

1. American Psychiatric Association. Diagnostic and Statistical Manual of Mental Disorders (DSM-5), 5th edn Washington, DC: American Psychiatric Association Publishing, 2013.

2. Lord, C., Elsabbagh, M., Baird, G., & Veenstra-Vanderweele, J. (2018). Autism spectrum disorder. Lancet (London, England), 392(10146), 508–520. 10.1016/S0140-6736(18)31129-2

3. Christensen, D. L., Braun, K. V. N., Baio, J., Bilder, D., Charles, J., Constantino, J. N., Daniels, J., Durkin, M. S., Fitzgerald, R. T., Kurzius-Spencer, M., Lee, L. C., Pettygrove, S., Robinson, C., Schulz, E., Wells, C., Wingate, M. S., Zahorodny, W., & Yeargin-Allsopp, M. (2018). Prevalence and Characteristics of Autism Spectrum Disorder Among Children Aged 8 Years - Autism and Developmental Disabilities Monitoring Network, 11 Sites, United States, 2012. Morbidity and mortality weekly report. Surveillance summaries (Washington, D.C.: 2002), 65(13), 1–23. 10.15585/mmwr.ss6513a1

4. Zeidan, J., Fombonne, E., Scorah, J., Ibrahim, A., Durkin, M. S., Saxena, S., Yusuf, A., Shih, A., & Elsabbagh, M. (2022). Global prevalence of autism: A systematic review update. Autism research: official journal of the International Society for Autism Research, 15(5), 778–790. 10.1002/aur.2696

5. Hossain, M. D., Ahmed, H. U., Jalal Uddin, M. M., Chowdhury, W. A., Iqbal, M. S., Kabir, R. I., Chowdhury, I. A., Aftab, A., Datta, P. G., Rabbani, G., Hossain, S. W., & Sarker, M. (2017). Autism Spectrum disorders (ASD) in South Asia: a systematic review. BMC psychiatry, 17(1), 281. 10.1186/s12888-017-1440-x

6. Chauhan, A., Sahu, J. K., Jaiswal, N., Kumar, K., Agarwal, A., Kaur, J., Singh, S., & Singh, M. (2019). Prevalence of autism spectrum disorder in Indian children: A systematic review and meta-analysis. Neurology India, 67(1), 100–104. 10.4103/0028-3886.253970

7. Doshi-Velez, F., Ge, Y., & Kohane, I. (2014). Comorbidity clusters in autism spectrum disorders: an electronic health record time-series analysis. Pediatrics, 133(1), e54–e63. 10.1542/peds.2013-0819

8. Chaste, P., & Leboyer, M. (2012). Autism risk factors: genes, environment, and gene-environment interactions. Dialogues in clinical neuroscience, 14(3), 281–292. 10.31887/DCNS.2012.14.3/pchaste

9. C Yuen, R. K., Merico, D., Bookman, M., L Howe, J., Thiruvahindrapuram, B., Patel, R. V., Whitney, J., Deflaux, N., Bingham, J., Wang, Z., Pellecchia, G., Buchanan, J. A., Walker, S., Marshall, C. R., Uddin, M., Zarrei, M., Deneault, E., D’Abate, L., Chan, A. J., Koyanagi, S., … Scherer, S. W. (2017). Whole genome sequencing resource identifies 18 new candidate genes for autism spectrum disorder. Nature neuroscience, 20(4), 602–611. 10.1038/nn.4524

10. Satterstrom, F. K., Kosmicki, J. A., Wang, J., Breen, M. S., De Rubeis, S., An, J. Y., Peng, M., Collins, R., Grove, J., Klei, L., Stevens, C., Reichert, J., Mulhern, M. S., Artomov, M., Gerges, S., Sheppard, B., Xu, X., Bhaduri, A., Norman, U., Brand, H., … Buxbaum, J. D. (2020). Large-Scale Exome Sequencing Study Implicates Both Developmental and Functional Changes in the Neurobiology of Autism. Cell, 180(3), 568–584.e23. 10.1016/j.cell.2019.12.036

11. Fu, J. M., Satterstrom, F. K., Peng, M., Brand, H., Collins, R. L., Dong, S., Wamsley, B., Klei, L., Wang, L., Hao, S. P., Stevens, C. R., Cusick, C., Babadi, M., Banks, E., Collins, B., Dodge, S., Gabriel, S. B., Gauthier, L., Lee, S. K., Liang, L., … Talkowski, M. E. (2022). Rare coding variation provides insight into the genetic architecture and phenotypic context of autism. Nature genetics, 54(9), 1320–1331. 10.1038/s41588-022-01104-0

12. Zhou, X., Feliciano, P., Shu, C., Wang, T., Astrovskaya, I., Hall, J. B., Obiajulu, J. U., Wright, J. R., Murali, S. C., Xu, S. X., Brueggeman, L., Thomas, T. R., Marchenko, O., Fleisch, C., Barns, S. D., Snyder, L. G., Han, B., Chang, T. S., Turner, T. N., Harvey, W. T., … Chung, W. K. (2022). Integrating de novo and inherited variants in 42,607 autism cases identifies mutations in new moderate-risk genes. Nature genetics, 54(9), 1305–1319. 10.1038/s41588-022-01148-2

13. Huguet, G., Ey, E., & Bourgeron, T. (2013). The genetic landscapes of autism spectrum disorders. Annual review of genomics and human genetics, 14, 191–213. 10.1146/annurev-genom-091212-153431

14. Wen, Y., Alshikho, M. J., & Herbert, M. R. (2016). Pathway Network Analyses for Autism Reveal Multisystem Involvement, Major Overlaps with Other Diseases and Convergence upon MAPK and Calcium Signaling. PloS one, 11(4), e0153329. 10.1371/journal.pone.0153329

15. Sandin, S., Lichtenstein, P., Kuja-Halkola, R., Hultman, C., Larsson, H., & Reichenberg, A. (2017). The Heritability of Autism Spectrum Disorder. JAMA, 318(12), 1182–1184. 10.1001/jama.2017.12141

16. Tick, B., Bolton, P., Happe, F., Rutter, M., and Rijsdijk, F. (2016). Heritability of autism spectrum disorders: a meta-analysis of twin studies. J. Child Psychol. Psychiatry 57, 585–595. 10.1111/jcpp.12499.

17. Hnoonual, A., Thammachote, W., Tim-Aroon, T., Rojnueangnit, K., Hansakunachai, T., Sombuntham, T., Roongpraiwan, R., Worachotekamjorn, J., Chuthapisith, J., Fucharoen, S., Wattanasirichaigoon, D., Ruangdaraganon, N., Limprasert, P., & Jinawath, N. (2017). Chromosomal microarray analysis in a cohort of underrepresented population identifies SERINC2 as a novel candidate gene for autism spectrum disorder. Scientific reports, 7(1), 12096. 10.1038/s41598-017-12317-3

18. Tammimies, K., Marshall, C. R., Walker, S., Kaur, G., Thiruvahindrapuram, B., Lionel, A. C., Yuen, R. K., Uddin, M., Roberts, W., Weksberg, R., Woodbury-Smith, M., Zwaigenbaum, L., Anagnostou, E., Wang, Z., Wei, J., Howe, J. L., Gazzellone, M. J., Lau, L., Sung, W. W., Whitten, K., … Fernandez, B. A. (2015). Molecular Diagnostic Yield of Chromosomal Microarray Analysis and Whole-Exome Sequencing in Children with Autism Spectrum Disorder. JAMA, 314(9), 895–903. 10.1001/jama.2015.10078

19. Pinto, D., Delaby, E., Merico, D., Barbosa, M., Merikangas, A., Klei, L., Thiruvahindrapuram, B., Xu, X., Ziman, R., Wang, Z., Vorstman, J. A., Thompson, A., Regan, R., Pilorge, M., Pellecchia, G., Pagnamenta, A. T., Oliveira, B., Marshall, C. R., Magalhaes, T. R., Lowe, J. K., … Scherer, S. W. (2014). Convergence of genes and cellular pathways dysregulated in autism spectrum disorders. American journal of human genetics, 94(5), 677–694. 10.1016/j.ajhg.2014.03.018

20. O’Roak, B. J., Vives, L., Girirajan, S., Karakoc, E., Krumm, N., Coe, B. P., Levy, R., Ko, A., Lee, C., Smith, J. D., Turner, E. H., Stanaway, I. B., Vernot, B., Malig, M., Baker, C., Reilly, B., Akey, J. M., Borenstein, E., Rieder, M. J., Nickerson, D. A., … Eichler, E. E. (2012). Sporadic autism exomes reveal a highly interconnected protein network of de novo mutations. Nature, 485(7397), 246–250. 10.1038/nature10989

21. O’Roak, B. J., Deriziotis, P., Lee, C., Vives, L., Schwartz, J. J., Girirajan, S., Karakoc, E., Mackenzie, A. P., Ng, S. B., Baker, C., Rieder, M. J., Nickerson, D. A., Bernier, R., Fisher, S. E., Shendure, J., & Eichler, E. E. (2011). Exome sequencing in sporadic autism spectrum disorders identifies severe de novo mutations. Nature genetics, 43(6), 585–589. 10.1038/ng.835

22. Bruel, A. L., Vitobello, A., Tran Mau-Them, F., Nambot, S., Sorlin, A., Denommé-Pichon, A. S., Delanne, J., Moutton, S., Callier, P., Duffourd, Y., Philippe, C., Faivre, L., & Thauvin-Robinet, C. (2020). Next-generation sequencing approaches and challenges in the diagnosis of developmental anomalies and intellectual disability. Clinical genetics, 98(5), 433–444. 10.1111/cge.13764

23. Bruno, L. P., Doddato, G., Valentino, F., Baldassarri, M., Tita, R., Fallerini, C., Bruttini, M., Lo Rizzo, C., Mencarelli, M. A., Mari, F., Pinto, A. M., Fava, F., Fabbiani, A., Lamacchia, V., Carrer, A., Caputo, V., Granata, S., Benetti, E., Zguro, K., Furini, S., … Ariani, F. (2021). New Candidates for Autism/Intellectual Disability Identified by Whole-Exome Sequencing. International journal of molecular sciences, 22(24), 13439. 10.3390/ijms222413439

24. Savatt, J. M., & Myers, S. M. (2021). Genetic Testing in Neurodevelopmental Disorders. Frontiers in pediatrics, 9, 526779. 10.3389/fped.2021.526779

25. Richards, S., Aziz, N., Bale, S., Bick, D., Das, S., Gastier-Foster, J., Grody, W. W., Hegde, M., Lyon, E., Spector, E., Voelkerding, K., Rehm, H. L., & ACMG Laboratory Quality Assurance Committee (2015). Standards and guidelines for the interpretation of sequence variants: a joint consensus recommendation of the American College of Medical Genetics and Genomics and the Association for Molecular Pathology. Genetics in medicine: official journal of the American College of Medical Genetics, 17(5), 405–424. 10.1038/gim.2015.30

26. World Health Organization. ICD-10: International Statistical Classification of Diseases and Related Health Problems: Tenth Revision, 2nd ed.; World Health Organization: Geneva, Switzerland, 2004.

27. Chakraborty, S., Bhatia, T., Sharma, V., Antony, N., Das, D., Sahu, S., Sharma, S., Shriharsh, V., Brar, J. S., Iyengar, S., Singh, R., Nimgaonkar, V. L., & Deshpande, S. N. (2021). Psychometric properties of a screening tool for autism in the community-The Indian Autism Screening Questionnaire (IASQ). PloS one, 16(4), e0249970. 10.1371/journal.pone.0249970

28. Javadi, A., Shamaei, M., Mohammadi Ziazi, L., Pourabdollah, M., Dorudinia, A., Seyedmehdi, S. M., & Karimi, S. (2014). Qualification study of two genomic DNA extraction methods in different clinical samples. Tanaffos, 13(4), 41–47.

29. Freed D. et al., The Sentieon Genomics Tools-A fast and accurate solution to variant calling from next-generation sequence data. BioRxiv:115717, 2017.

30. https://cloud.google.com/life-sciences/docs/tutorials/deepvariant

31. Landrum, M. J., Lee, J. M., Benson, M., Brown, G., Chao, C., Chitipiralla, S., Gu, B., Hart, J., Hoffman, D., Hoover, J., Jang, W., Katz, K., Ovetsky, M., Riley, G., Sethi, A., Tully, R., Villamarin-Salomon, R., Rubinstein, W., & Maglott, D. R. (2016). ClinVar: public archive of interpretations of clinically relevant variants. Nucleic acids research, 44(D1), D862–D868. 10.1093/nar/gkv1222

32. Welter, D., MacArthur, J., Morales, J., Burdett, T., Hall, P., Junkins, H., Klemm, A., Flicek, P., Manolio, T., Hindorff, L., & Parkinson, H. (2014). The NHGRI GWAS Catalog, a curated resource of SNP-trait associations. Nucleic acids research, 42(Database issue), D1001–D1006. 10.1093/nar/gkt1229

33. 1000 Genomes Project Consortium, Auton, A., Brooks, L. D., Durbin, R. M., Garrison, E. P., Kang, H. M., Korbel, J. O., Marchini, J. L., McCarthy, S., McVean, G. A., & Abecasis, G. R. (2015). A global reference for human genetic variation. Nature, 526(7571), 68–74. 10.1038/nature15393

34. Conti, V., Carabalona, A., Pallesi-Pocachard, E., Parrini, E., Leventer, R. J., Buhler, E., McGillivray, G., Michel, F. J., Striano, P., Mei, D., Watrin, F., Lise, S., Pagnamenta, A. T., Taylor, J. C., Kini, U., Clayton-Smith, J., Novara, F., Zuffardi, O., Dobyns, W. B., Scheffer, I. E., … Guerrini, R. (2013). Periventricular heterotopia in 6q terminal deletion syndrome: role of the C6orf70 gene. Brain: a journal of neurology, 136(Pt 11), 3378–3394. 10.1093/brain/awt249

35. Rivière, J. B., Ramalingam, S., Lavastre, V., Shekarabi, M., Holbert, S., Lafontaine, J., Srour, M., Merner, N., Rochefort, D., Hince, P., Gaudet, R., Mes-Masson, A. M., Baets, J., Houlden, H., Brais, B., Nicholson, G. A., Van Esch, H., Nafissi, S., De Jonghe, P., Reilly, M. M., … Rouleau, G. A. (2011). KIF1A, an axonal transporter of synaptic vesicles, is mutated in hereditary sensory and autonomic neuropathy type 2. American journal of human genetics, 89(2), 219–230. 10.1016/j.ajhg.2011.06.013

36. Klebe, S., Lossos, A., Azzedine, H., Mundwiller, E., Sheffer, R., Gaussen, M., Marelli, C., Nawara, M., Carpentier, W., Meyer, V., Rastetter, A., Martin, E., Bouteiller, D., Orlando, L., Gyapay, G., El-Hachimi, K. H., Zimmerman, B., Gamliel, M., Misk, A., Lerer, I., … Stevanin, G. (2012). KIF1A missense mutations in SPG30, an autosomal recessive spastic paraplegia: distinct phenotypes according to the nature of the mutations. European journal of human genetics: EJHG, 20(6), 645–649. 10.1038/ejhg.2011.261

37. Lee, J. R., Srour, M., Kim, D., Hamdan, F. F., Lim, S. H., Brunel-Guitton, C., Décarie, J. C., Rossignol, E., Mitchell, G. A., Schreiber, A., Moran, R., Van Haren, K., Richardson, R., Nicolai, J., Oberndorff, K. M., Wagner, J. D., Boycott, K. M., Rahikkala, E., Junna, N., Tyynismaa, H., … Michaud, J. L. (2015). De novo mutations in the motor domain of KIF1A cause cognitive impairment, spastic paraparesis, axonal neuropathy, and cerebellar atrophy. Human mutation, 36(1), 69– 78. 10.1002/humu.22709

38. Tomaselli, P. J., Rossor, A. M., Horga, A., Laura, M., Blake, J. C., Houlden, H., & Reilly, M. M. (2017). A de novo dominant mutation in KIF1A associated with axonal neuropathy, spasticity and autism spectrum disorder. Journal of the peripheral nervous system: JPNS, 22(4), 460–463. 10.1111/jns.12235

39. Kellogg, G., Sum, J., & Wallerstein, R. (2013). Deletion of 3p25.3 in a patient with intellectual disability and dysmorphic features with further definition of a critical region. American journal of medical genetics. Part A, 161A(6), 1405–1408. 10.1002/ajmg.a.35876

40. Sessa, A., Fagnocchi, L., Mastrototaro, G., Massimino, L., Zaghi, M., Indrigo, M., Cattaneo, S., Martini, D., Gabellini, C., Pucci, C., Fasciani, A., Belli, R., Taverna, S., Andreazzoli, M., Zippo, A., & Broccoli, V. (2019). SETD5 Regulates Chromatin Methylation State and Preserves Global Transcriptional Fidelity during Brain Development and Neuronal Wiring. Neuron, 104(2), 271–289.e13. 10.1016/j.neuron.2019.07.013

41. Chen, Q., Sun, Z., Li, J., Zhang, D., Guo, B., & Zhang, T. (2021). SET Domain-Containing Protein 5 Enhances the Cell Stemness of Non-Small Cell Lung Cancer via the PI3K/Akt/mTOR Pathway. Journal of environmental pathology, toxicology and oncology: official organ of the International Society for Environmental Toxicology and Cancer, 40(2), 5563. 10.1615/JEnvironPatholToxicolOncol.2021036991

42. Jethmalani, Y., Tran, K., Negesse, M. Y., Sun, W., Ramos, M., Jaiswal, D., Jezek, M., Amos, S., Garcia, E. J., Park, D., & Green, E. M. (2021). Set4 regulates stress response genes and coordinates histone deacetylases within yeast subtelomeres. Life science alliance, 4(12), e202101126. 10.26508/lsa.202101126

43. Wagner, E. J., & Carpenter, P. B. (2012). Understanding the language of Lys36 methylation at histone H3. Nature reviews. Molecular cell biology, 13(2), 115–126. 10.1038/nrm3274

44. Grozeva, D., Carss, K., Spasic-Boskovic, O., Parker, M. J., Archer, H., Firth, H. V., Park, S. M., Canham, N., Holder, S. E., Wilson, M., Hackett, A., Field, M., Floyd, J. A., UK10K Consortium, Hurles, M., & Raymond, F. L. (2014). De novo loss-of-function mutations in SETD5, encoding a methyltransferase in a 3p25 microdeletion syndrome critical region, cause intellectual disability. American journal of human genetics, 94(4), 618–624. 10.1016/j.ajhg.2014.03.006

45. Iwasawa, S., Yanagi, K., Kikuchi, A., Kobayashi, Y., Haginoya, K., Matsumoto, H., Kurosawa, K., Ochiai, M., Sakai, Y., Fujita, A., Miyake, N., Niihori, T., Shirota, M., Funayama, R., Nonoyama, S., Ohga, S., Kawame, H., Nakayama, K., Aoki, Y., Matsumoto, N., … Kure, S. (2019). Recurrent de novo MAPK8IP3 variants cause neurological phenotypes. Annals of neurology, 85(6), 927–933. 10.1002/ana.25481

46. Platzer, K., Sticht, H., Edwards, S. L., Allen, W., Angione, K. M., Bonati, M. T., Brasington, C., Cho, M. T., Demmer, L. A., Falik-Zaccai, T., Gamble, C. N., Hellenbroich, Y., Iascone, M., Kok, F., Mahida, S., Mandel, H., Marquardt, T., McWalter, K., Panis, B., Pepler, A., … Jamra, R. (2019). De Novo Variants in MAPK8IP3 Cause Intellectual Disability with Variable Brain Anomalies. American journal of human genetics, 104(2), 203–212. 10.1016/j.ajhg.2018.12.008

